# Plasma water T_2_ detects age-stratified differences in cardiometabolic health among familial CCM patients with Hispanic CCM1 mutation

**DOI:** 10.1101/2023.08.10.23293944

**Authors:** Jacob Croft, Diana F. Sandoval, David Cistola, Jun Zhang

## Abstract

**Introduction:** Cerebral cavernous malformations (CCMs) are abnormal clusters of capillaries in the nervous system. This pilot study analyzed the cardiometabolic health status of individuals with familial CCMs caused by a rare mutation in the *CCM1* gene (fCCM1). The aim was to compare plasma water T_2_ values from individuals with fCCM1 with values from metabolically unhealthy and healthy individuals with no known CCM mutations.

**Design:** This observational, cross-sectional study included 75 participants: 11 fCCM1 patients, 24 metabolically unhealthy and 40 metabolically healthy individuals. Plasma water T_2_, an early, global and practical marker of cardiometabolic health, was measured in the time domain using benchtop magnetic resonance relaxometry. The results were stratified by age (≤ 45 vs. >45 years). Group means were compared using Welch’s one-way ANOVA and *post hoc* Tukey-Kramer tests. Multivariable linear regression, with T_2_ as the outcome variable, was used to explore associations with age, gender, Hispanic ethnicity and fCCM1 status.

**Results:** In the younger age stratum, the fCCM1 group had a mean plasma water T_2_ value comparable to the metabolically healthy group (p=0.6388), but higher than the unhealthy group (p<0.0001). By contrast, in the older stratum, the mean plasma water T_2_ value for the fCCM1 group was comparable to the metabolically *un*healthy group (p=0.7819) and lower than the healthy group (p=0.0005). Multivariable linear regression revealed that age and the *interaction* between age and fCCM1 status were significant predictors of T_2_, even after adjusting for gender and Hispanic ethnicity.

**Conclusion:** Plasma water T_2_ shows potential as a biomarker for assessing the health status of individuals with fCCM1. Further research is needed to validate these preliminary observations and elucidate the association between CCMs and cardiometabolic health.

## INTRODUCTION

Cerebral cavernous malformations (CCMs) are vascular lesions characterized by abnormal clusters of capillaries found in both the central and peripheral nervous system. The development of CCM lesions is attributed to endothelial cells that are dysfunctional, exhibiting diminished cellular adhesion. This condition leads to heightened vascular permeability and an increased susceptibility to hemorrhage. While relatively uncommon, CCMs account for approximately 15% of all vascular malformations, despite occurring in only approximately 0.5% of the population [1-3]. CCMs are characterized by two distinct features, each associated with both common and unique causative genetic mutation landscapes. These features encompass familial CCMs (fCCMs), which are inherited, and sporadic CCMs (sCCMs), which arise de novo [2, 4, 5]. The focus of this study is on a single “common Hispanic” fCCM1 mutation due to its inheritable nature and traceable genetic lineage route.

The majority of fCCMs are attributed to mutations in three genes: CCM1 (KRIT1), CCM2 (MGC4607), and CCM3 (PDCD10) [1-3]. Among these mutations, CCM1 loss-of-function (LOF) is the most common, accounting for approximately 50% of all causative mutations [1-3]. These mutations result in vascular malformations that may present with symptoms in some patients or go undetected in asymptomatic individuals. Clinical manifestations of CCMs include headaches/migraines, focal neural deficits, seizures, hemorrhage, and stroke [1-3]. The incomplete penetrance exhibited by these mutations poses challenges for early detection in clinical settings. Currently, detection and diagnosis rely on methods such as magnetic resonance imaging (MRI), gradient-echo (GRE), susceptibility-weighted imaging (SWI), or pathological analysis of biopsied tissue [6, 7]. However, these diagnostic tools have practical limitations for ongoing monitoring due to cost and accessibility, making it necessary to find accurate and affordable methods to assess the risk of acute hemorrhagic events.

The loss-of-function (LOF) of CCM1 leads to an increased vulnerability to oxidative DNA damage, which in turn activates DNA damage sensors, repair genes, and triggers an apoptotic response. Based on these observations, it suggests that CCM1 plays a vital role in preserving the intracellular balance of reactive oxygen species (ROS), thus preventing cellular dysfunctions caused by ROS [8]. Moreover, CCM1 functions to inhibit the upregulation of c-Jun induced by oxidative stimuli [9] and prevents abnormal activation of the Nrf2 stress defense system, along with its downstream effectors HO-1 and Glo1 [10]. The effect of CCM1 deficiency on the levels of reactive oxygen species (ROS) and redox status has been documented in multi-omics data [11, 12].

The LOF of CCM1 not only affects the process of angiogenesis, but also influences several other vital processes including metabolic syndrome (MetS). The LOF of CCM1 has been shown to impact food intake, systemic glucose disposal as well as plasma insulin levels [13]. In addition to these impaired processes, it has been found that LOF of CCM1 has been linked to higher levels of blood pressure and higher fasting glucose levels, which increase the intensity of symptoms of vascular malformations [14].

A recently developed technique for the early detection of the underlying pathophysiology of metabolic syndrome utilizes the plasma water transverse relaxation time constant (T_2_), measured using benchtop magnetic resonance relaxometry [15-18]. Conceivably, this early detection tool could assess the risk for, or monitor the status of, hemorrhagic events to allow the patient-clinician team to preemptively treat disorders known to increase symptoms. Therefore, the aim of this initial study was to compare plasma water T_2_ values for individuals with a known CCM1 mutation with those of metabolically healthy and unhealthy controls with no known CCM mutations.

## EXPERIMENTAL METHODS

### Sample Recruitment

We recruited participants from a well-established patient cohort of familial CCM (fCCM) caused by the "common Hispanic CCM1 mutation" from a large Baca family pedigree [19, 20]. All CCM patients (N=11) carried the same Baca CCM1 hemizygous mutation. The participants recruited from the CCM cohort had experienced hemorrhagic lesions in close proximity to the time of sample collection. All CCM1 participants in this study fell within the age range of 16-61, in compliance with the protocols set by the local Institutional Review Board (IRB-E21010).

Individuals with no known CCM mutations (“controls”) who were metabolically healthy (n=40) or metabolically unhealthy (n=24). Of the blood samples obtained from the 64 control subjects with no known CCM mutation, 20 of the samples were de-identified and provided by the Regional Laboratory of the University Medical Center of El Paso (IRB-E21010). The remaining 44 samples were from Phase 2 participants in the Fort Worth T_2_ Study, as described elsewhere [15, 16]. That study was approved by the Institutional Review Board of the University of North Texas Health Science Center in Fort Worth. Each of the 44 participants was metabolically phenotyped with a medical history and physical exam, including ∼130 clinical lab tests and biomarkers. These individuals were asymptomatic, with no active acute or chronic disease, and no history of blood disorders or hemorrhagic stroke. The classification of metabolically healthy vs. unhealthy was based on the ROC-calibrated cut point for plasma water T_2_ (700 msec) in samples that had undergone one freeze-thaw cycle [16].

### Benchtop Time-Domain Magnetic Resonance (MR) Relaxometry of Human Plasma

#### MR relaxometry

In contrast to frequency-domain MR imaging (MRI) and spectroscopy, MR relaxometry records and analyzes data directly in the time domain without Fourier transformation. The key advantage of MR relaxometry is simplicity and efficiency: it can be performed with a benchtop, compact or miniaturized device containing a low-field permanent magnet. Unlike spectroscopy and imaging, MR relaxometry measurements can be made using compact or portable devices at the point of care.

The plasma water T_2_ value is highly sensitive to the rotational/translational diffusion of water molecules as they bind and exchange on and off abundant proteins and lipoproteins. Metabolic shifts in the numbers and sizes of proteins and lipoprotein particles occur as a result of insulin resistance, inflammation (acute phase response), oxidative stress and the pro-coagulation state, resulting in a net decrease in T_2_. Thus, plasma water T_2_ provides an overall or global assessment of cardiometabolic health and detects the underlying pathophysiology of metabolic syndrome. In prior work, we have effectively employed compact MR relaxometry to examine human blood samples and extract significant health information [15-18, 21-23].

#### Sample preparation

The current study utilized bio-banked human plasma samples frozen once at -80°C. Previous research comparing freshly drawn samples with plasma and serum samples that had undergone a single freeze-thaw cycle showed that the health information associated with T_2_ was preserved in the frozen samples [16]. Here, plasma samples were retrieved from the -80°C freezer one day prior to the measurement and gradually thawed overnight at 4°C. On the following day, approximately 40-50 μL of unmodified human plasma, corresponding to a sample height of 1 cm, was carefully transferred into a 3 mm coaxial insert (catalog NE-10-CIC-SB) placed inside an empty outer 10 mm NMR tube (NE-L10-7, New Era Enterprises, Inc, Vineland, NJ). Prior to MR relaxometry analysis, the plasma samples were pre-incubated at 37°C for 30 minutes.

#### Data acquisition

Plasma water ^1^H T_2_ relaxation decay curves were measured in the time domain at 37°C using a Bruker mq20-NF Minispec instrument operating at 0.47 Tesla (20 MHz for ^1^H). The instrument was equipped with a H20-10-25AVGX4 probe assembly housed within the magnet’s 25 mm air gap. A modified Carr-Purcell-Meiboom Gill (CPMG) pulse sequence, including a repeated (τ-180°-τ) pulse train, was employed with data acquisition in the middle of every other 2τ delay [15, 21]. To minimize the influence of translational diffusion, a short 2τ delay time of 0.38 msec was utilized to ensure an adequate baseline for multi-exponential analysis, the spin-echo train was recorded for a minimum of 8 times T_2_. Each experiment recorded 16 signal-averaged decay curves. The recording time for each experiment was approximately 4 minutes, and each sample was measured three times to assess precision. Before each round of the pulse sequence, a relaxation delay of 8 times T_1_ (longitudinal or spin-lattice relaxation time) was implemented to allow the system to return to thermal equilibrium. To estimate T_1_, an inversion-recovery pulse sequence with a series of 20 time delays was employed.

#### Data processing and analysis

The deconvolution of each raw multi-exponential CPMG decay curve was performed using a discrete inverse Laplace transformation algorithm. This algorithm is implemented in XpFit software (Soft Science, Alango, Ltd.). This algorithm permits the user to specify a fixed number of exponential components (here, 3), which is crucial for ensuring meaningful subject-to-subject comparison of T_2_ values [15, 16, 21]. The discrete inverse Laplace transform algorithm yields a T_2_ profile represented as signal intensity (y-axis) vs. T_2_ (x-axis). This profile is not to be confused with a MR spectrum, where the x-axis measures frequency. The discrete inverse Laplace transform effectively resolves the water T_2_ hydrogen peak (spike) from the peaks attributed to non-water hydrogens from proteins, lipids and metabolites. The reported plasma water T_2_ values represent the mean of three repeats and were highly reproducible (SEM ∼1%).

### Statistical Analysis of the Human Dataset

Multiple mean comparisons and multivariable linear regression analyses were performed using RStudio v. 2023.3.0.386 [24] and JMP Pro v. 16.2 (SAS Institute, Cary, NC, USA). Mean comparisons utilized Welch’s one-way ANOVA and *post hoc* Tukey-Cramer tests. The purpose was to compare the mean plasma water T_2_ value for the fCCM1 group to those for healthy and metabolically unhealthy control groups. Linear regression was performed to identify models that optimized the prediction of mean plasma water T_2_ (outcome variable). The candidate predictors were fCCM1 status (known carrier of the Baca CCM1 hemizygous mutation) or no known CCM mutations, age (continuous or categorical), sex (gender at birth) and Hispanic vs. non-Hispanic ethnic status.

## RESULTS

The characteristics of the study population (n=75) are summarized in Table 1. Overall, the study population had higher numbers of female vs. male participants, especially in the fCCM1 group. The age range was 16-61. The median age for the fCCM1 and unhealthy groups were similar: 44 and 47 years of age, respectively. The median plasma water T_2_ value for the fCCM1 group (695.1 msec) fell between those of the unhealthy (661.8 msec) and healthy (754.7 msec) control groups. Healthy and unhealthy were defined using a T_2_ cut point of 700.1 msec, calibrated using receiver operator characteristic (ROC) curves, where poor cardiometabolic health (“unhealthy”) was defined as a combination of hyperinsulinemia, dyslipidemia and inflammation [16].

**Table 1.** Characteristics of the Study Population (n=75).

| **Group** | **Number** | **Sex<sup>1</sup>** | **Hispanic Status<sup>2</sup>** | **Age<sup>3</sup>** | **Plasma water T<sub>2</sub><sup>2</sup>** |
| --- | --- | --- | --- | --- | --- |
| fCCM1<sup>4</sup> | 11 | 9 F, 2 M | 10 H, 1 NH | 44 (15), 16-61 | 695.1 (81.3), 515.2-767.8 |
| unhealthy<sup>5</sup> | 24 | 16 F, 8 M | 11H, 13 NH | 47 (25), 19-61 | 661.8 (35.5), 603.7-695.7 |
| healthy<sup>6</sup> | 40 | 20 F, 20 M | 10 H, 30 NH | 30 (23), 16-61 | 754.7 (55.9), 700.1-840.4 |
| all non-fCCM1<sup>7</sup> | 64 | 36 F, 28 M | 31 H, 44 NH | 37 (25), 16-61 | 719.8 (91.9), 603.7-840.4 |

<sup>1</sup>F, female; M, male
<sup>2</sup>H, Hispanic; NH, non-Hispanic
<sup>3</sup>median (interquartile range), full range
<sup>4</sup>individuals carrying the same Baca familial CCM1 hemizygous mutation
<sup>5</sup>individuals with no known CCM mutation and poor cardiometabolic health (T<sub>2</sub> ≤700.1 msec)
<sup>6</sup>individuals with no known CCM mutation and good cardiometabolic health (T<sub>2</sub> >700.1 msec)
<sup>7</sup>individuals with no known CCM mutation; combination of unhealthy and healthy groups

Figure 1 shows plasma water T_2_ values for three categories of health status, stratified by age. The horizontal black line indicates the robust mean plasma water T_2_ value for each category. The upper and lower ends of each colored box delineate the interquartile range (IQR), and the whiskers cover the full range of observations. Each participant is represented by solid black dots, and the dots are horizontally staggered to minimize overlap and aid in visualization. The red boxes represent fCCM1, and the green and blue boxes represent healthy and unhealthy controls without known CCM mutations, respectively.

**Figure 1.**
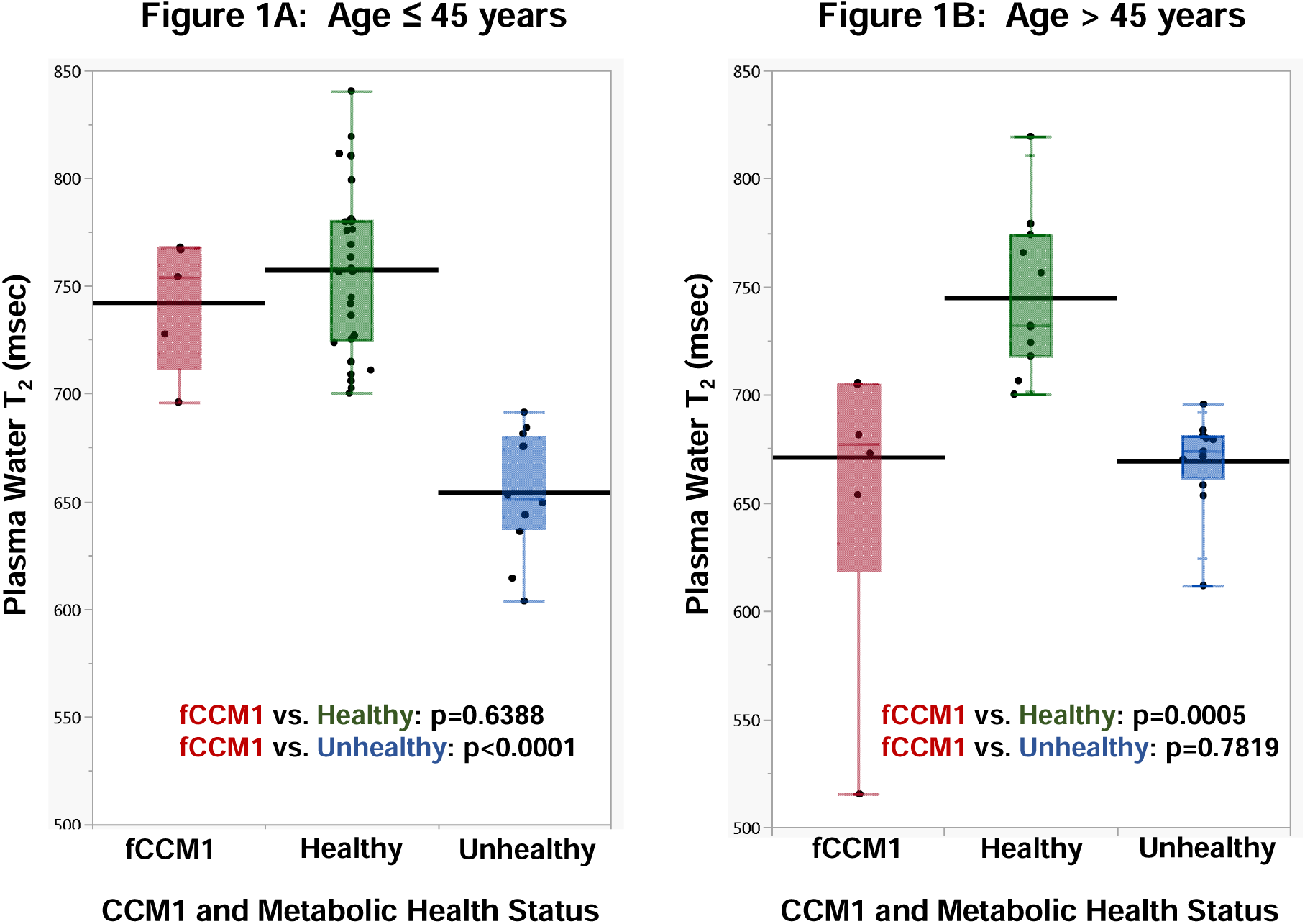
Box plots for plasma water T_2_ vs. CCM1 and cardiometabolic health status category; each participant is represented by a solid black The lower and upper bounds of each colored box represent the 25^th^ and 75^th^ percentiles, respectively, and the horizontal colored line within each box, the 50^th^ percentile; red, participants carrying the same fCCM1 mutation; green, control (non-fCCM1) participants with good cardiometabolic health, (T_2_ >700.1 msec); blue, control participants with poor cardiometabolic health (T_2_ ≤ 700.1 msec); abbreviations: fCCM1, cerebral cavernous malformation gene 1 mutation; T_2_, transverse or spin-spin relaxation time from magnetic resonance relaxometry; msec, milliseconds.

Using Welch’s one-way ANOVA and post hoc Tukey-Kramer tests, the mean plasma water T_2_ values were compared. As shown in Fig. 1A (younger age stratum), the robust mean T_2_ value for the fCCM1 group was 742.4 msec. It was statistically indistinguishable from that for healthy group (757.6 msec; p=0.6388), but different from the metabolically unhealthy group (654.2 msec; p<0.0001). By contrast, for the older age stratum (Fig. 1B), the mean plasma water T_2_ value for the fCCM1 group was 671.0 msec. It was statistically indistinguishable from the metabolically *un*healthy group (669.3 msec; p=0.7819), but different from the healthy group (745.0 msec; p=0.0005). Based on a previously published cut point for samples that underwent one freeze-thaw cycle, plasma water T_2_ values ≤700.1 msec are consistent with poor cardiometabolic health, i.e., hyperinsulinemia plus dyslipidemia plus inflammation [16].

This stratified analysis suggested that age may modify the association between plasma water T_2_ and fCCM1 status. To further explore the potential interaction between age and fCCM1 status, two multiple linear regression models were generated. The outcome variable was plasma water T_2_, and the predictors were sex, Hispanic ethnicity, age, fCCM1 status and the interaction between age and fCCM1 status.

The regression parameters for Model 1 are displayed in Table 2A. Shown are the beta coefficients (“Estimate” column) and p-values (“Prob>|t|” column) when age was analyzed as a continuous variable. In this model, sex and Hispanic ethnicity were not significant predictors of plasma water T_2_. Rather, the statistically significant predictors were age and the interaction term for age*fCCM1 status.

**Table 2.**
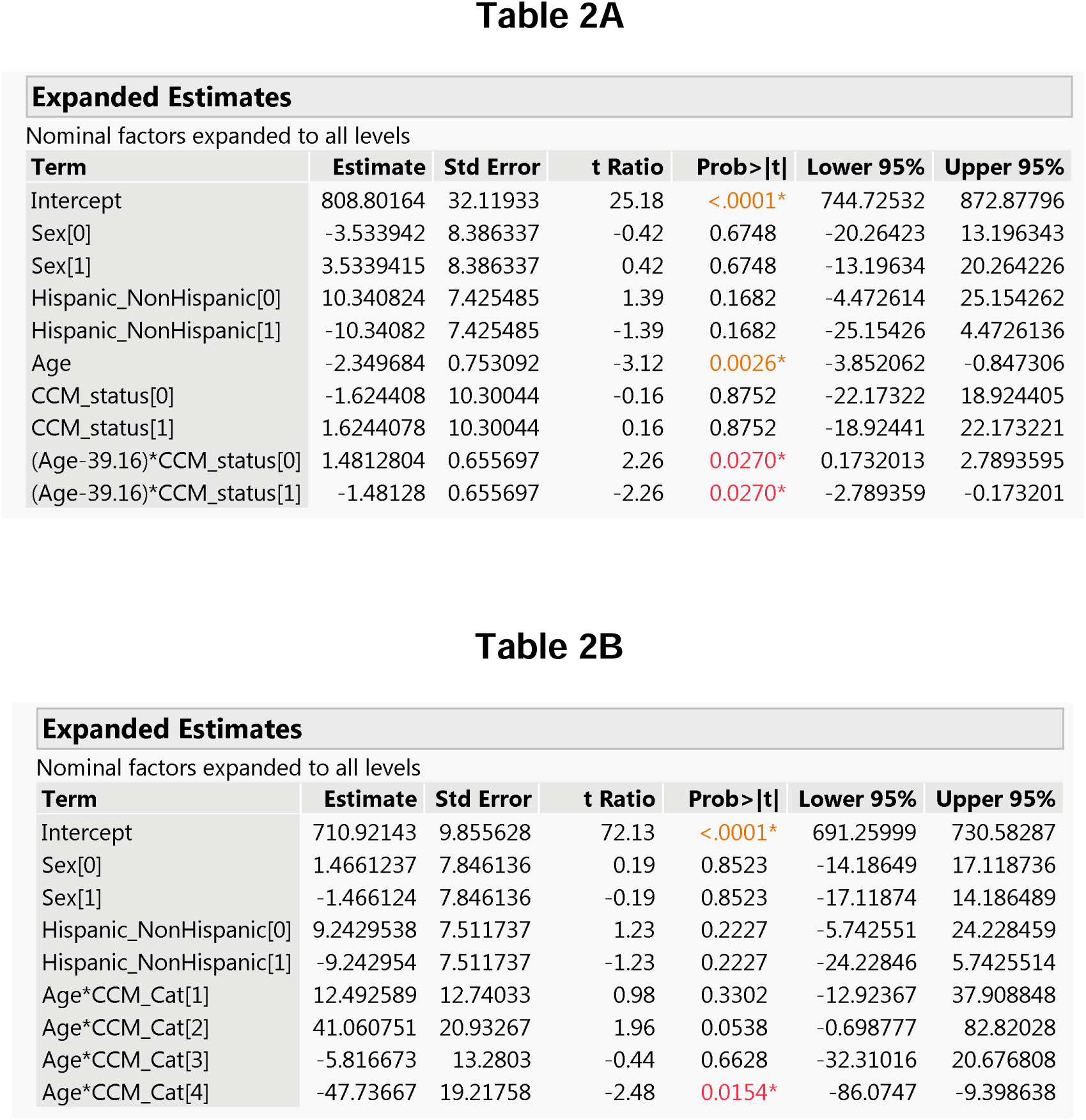
Estimates of beta-coefficients from multivariable linear regression models that include sex, Hispanic ethnicity status and either: (A) age as a continuous variable, along with an interacting variable for age*CCM category; or (B) age as part of a 4-level categorical interacting variable that also includes CCM status. Regression analysis was performed in JMP v16.2, which uses effect coding for categorical variables. <u>Abbreviations</u>: Prob>|t|, p-value at α=0.05; Sex[0]=male; Sex[1]=female; Hispanic_ NonHispanic[0]= not Hispanic; Hispanic_NonHispanic[1] = Hispanic ethnicity; CCM_status[0]=no known CCM mutation; CCM_status[1]=carries fCCM1 mutation; Age*CCM_Cat[1]=Age ≤45 & no known CCM1 mutation; Age*CCM_Cat[2]=Age ≤45 & fCCM1 mutation; Age*CCM_Cat[3]=Age >45 & no known CCM mutation; Age*CCM1_Cat[4]=Age >45 & fCCM1 mutation.

The interaction between continuous age and CCM1 status can be visualized using an interaction plot, as derived from Model 1. Figure 2 shows two regression lines for plasma water T_2_ vs. age: one for individuals with fCCM1 (1, red line) and the other for those without known CCM mutations (0, black line). The slope for fCCM1 individuals was steeper than that for controls with no known CCM mutations. This change in slope provides further evidence for an interaction (effect modification) between age and fCCM1 mutation status. Stated another way, fCCM1 status modifies or *magnifies* the association between plasma water T_2_ and age.

**Figure 2.**
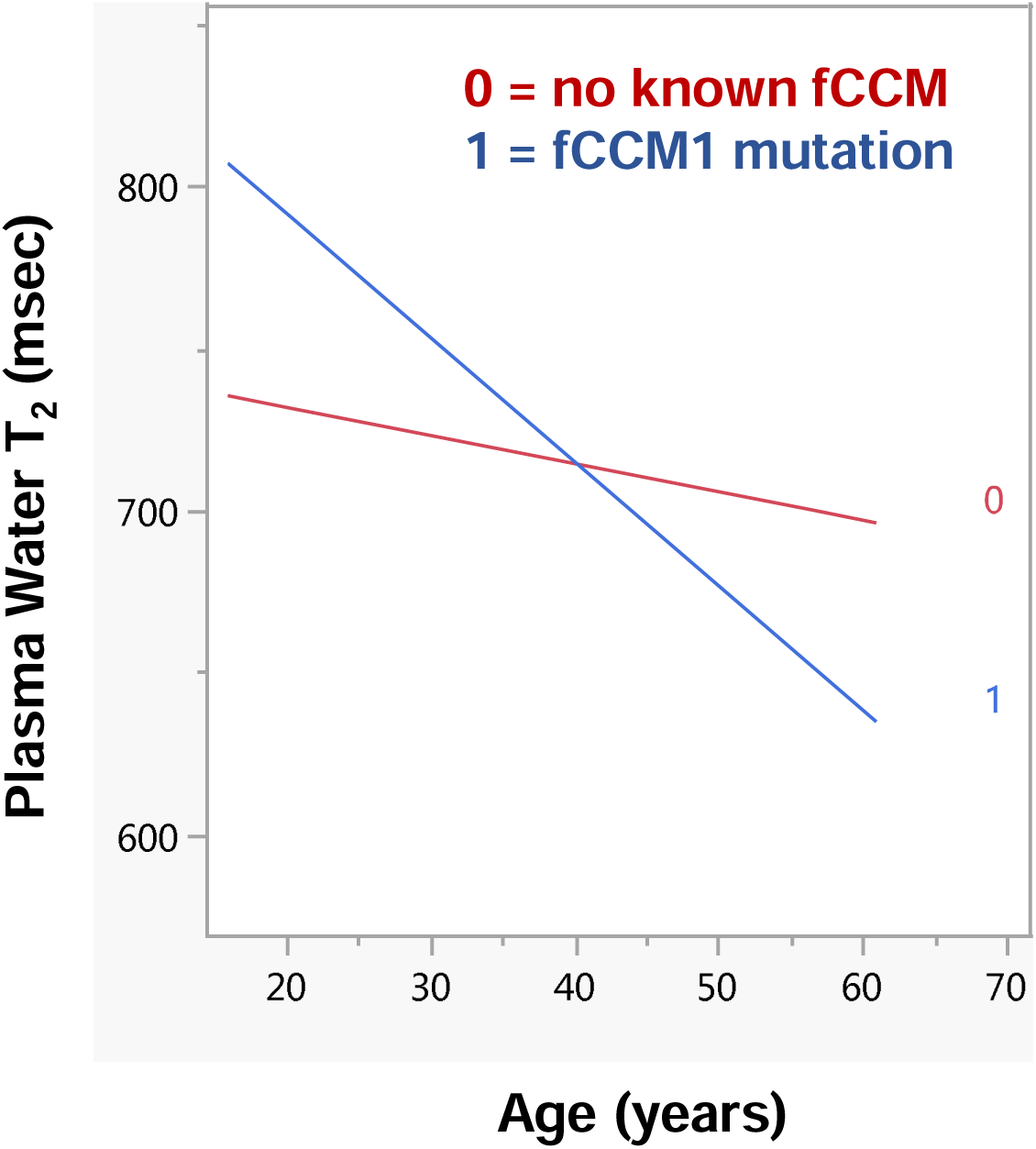
Interaction plot showing the modification of the slope of plasma water T_2_ vs. age, by fCCM1 gene status; 1 or red, regression line for individuals carrying the fCCM1 mutation; 0 or black, regression line for participants with no known fCCM mutations.

Table 2B lists the beta coefficients and p-values for Model 2: similar to Model 1, except that age was categorical and the interaction between age and CCM1 status was coded as a categorical variable with 4 levels. Level 1 represented individuals with age ≤45 and no known CCM mutations. Level 2 represented individuals with age ≤45 and fCCM1. Level 3 represented age >45 and no known CCM mutations. Level 4 represents age >45 and fCCM1. As shown in the bottom row of Table 2B, the *combination* between age >45 and fCCM1 (Level 4) was a significant predictor of plasma water T_2_ (p=0.0154). None of the other levels or categorical predictors were statistically significant. Note that Level 2, while insignificant (p=0.0533), was close to the α=0.05 significance threshold.

## DISCUSSION

The results from this study revealed a distinct age-stratified difference in mean plasma water T_2_ values for individuals carrying the Baca familial “common Hispanic” hemizygous mutation in the *CCM1* gene. Younger fCCM1 participants (≤45 years of age) had a mean T_2_ value consistent with good cardiometabolic health. By contrast, fCCM1 participants >45 years of age exhibited a mean T_2_ value consistent with poor cardiometabolic health. Subsequent regression analysis indicated that the association between plasma water T_2_ and fCCM1 status was modified by age. Likewise, the association between plasma water T_2_ and age was modified by fCCM1 status, even after adjusting for sex and Hispanic ethnicity. This effect modification manifested itself in the one-way ANOVA plots stratified by age, along with the statistically significant interaction terms in two multiple regression models.

The discrete decline in plasma water T_2_ values in middle age aligns with the average known onset of CCM lesions occurring between the third and fifth decade of life [25]. The incomplete penetrance of CCMs suggests that an additional trigger, whether environmental or genetic, is required for the progression of vascular lesions towards hemorrhagic events. Several potential triggers have been proposed, but their validity is still debated due to conflicting results [26]. These triggers include factors of both pathological and physiological nature, such as the gut microbiota theory [27], the anticoagulant vascular domain theory [28], levels of antioxidants like vitamin D [29], retinoic acid levels [30] and perturbed progesterone (PRG) signaling [26].

Interestingly, the last three factors are all associated with signal pathways in breast cancer nuclear receptor categories, indicating a potential connection among them. The progesterone (PRG) pathway is of particular importance in the context of CCMs, as the CCM signaling complex (CSC) has been found to involve both classic nuclear PRG receptors (nPR) and non-classic membrane PRG receptors (mPR), forming the CmPn (CSC-mPR-PRG-nPR) signal network that plays a role in the development and maintenance of the blood-brain barrier [31]. CCM patients exhibit varying PRG levels throughout their lives, and CCM mutations can lead to metabolic dysfunction and decreased plasma glucose levels, as demonstrated in studies involving Ccm mutant mice [13]. The mouse studies have been independently validated in a recent human retrospective study [14].

Plasma water T_2_ values have shown high sensitivity in detecting metabolic abnormalities, particularly insulin resistance measured as the combination of hyperinsulinemia, dyslipidemia and inflammation [16]. Although the current study was preliminary, we observed that T_2_ values for individuals with fCCM1 abruptly declined in mid-life. This decline was more abrupt than observed for healthy and unhealthy individuals without known fCCM mutations. Plasma water T_2_ could conceivably detect the trigger-point change in health status in fCCM1 patients. Further investigating the association between CCMs and plasma water T_2_ values presents an opportunity to develop a prognostic biomarker that could aid in detecting changes in cardiometabolic health and possibly, the onset or worsening of vascular malformations as precursors of hemorrhagic stroke.

The interaction between fCCM1 status and age in its impact on cardiometabolic health, as observed here, is consistent with the results of a clinical retrospective study that reported a higher prevalence of MetS in patients aged 50 and above [14]. Furthermore, the same study revealed that Mexican-Hispanic CCM subjects who developed MetS had a higher susceptibility to hemorrhagic stroke [14]. Currently, blood tests are capable of detecting the occurrence of *ischemic* strokes. However, there is a need for the development of a prognostic biomarker for the early prediction and prevention of *hemorrhagic* strokes.

### Limitations

This pilot study had several significant limitations. First, the study sample size for the fCCM1 group was small: 11 individuals. Mutations in the *CCM1* gene are rare; identifying and recruiting such individuals and obtaining blood samples is challenging. Second, the sex of the fCCM1 group was skewed, as 9 of the 11 participants were female. Older women tend to exhibit a stronger response to changes in progesterone compared to men, which could have potentially contributed to disruptions in these pathways and their effects on the blood-brain barrier. Sex-related differences should be taken into consideration when interpreting the study’s findings and understanding their broader applicability. Third, there was a lack of individuals in the fCCM1 group in the 20 to 35 age range, even though the full age range for that group was 16-61. Caution should be used in applying these results to fCCM1 individuals in their prime childbearing years. Fourth, the data provided with the fCCM1 samples, and 20 of the 64 control samples, were limited to sex, age and Hispanic status, with no access to medical histories, health records, or other test results. This precluded a broader assessment of health status and limited the statistical analyses that could be performed. Fifth, the fCCM1 mutation studied here was identified in a Hispanic extended family living in the southwestern border region of the U.S. Although the representation of ethnicities in the study are skewed, it is recognized that there is a higher risk of CCM inheritance among Hispanic families [32, 33]. There were insufficient numbers of individuals in other ethnic or racial groups to assess the impact of those variables on fCCM1 status or health outcomes. Sixth, the non-fCCM1 individuals were not genotyped, although it is highly unlikely that any carried rare CCM gene mutations. Finally, as a pilot study focused on a rare gene mutation, the results from this study are not generalizable to other populations.

### Strengths and Significance

In spite of the limitations listed above, the results from this pilot study revealed statistically significant age-stratified differences in cardiometabolic health, as assessed by plasma water T_2_, among fCCM1 patients. The findings provide justification for a larger study with more health measurements to validate and extend these findings. There is an unmet need for an affordable, easily accessible test for monitoring the onset and progression of CCM lesions, monitoring the decline cardiometabolic health, and predicting the risk of hemorrhagic stroke. Although further research and validation is needed, plasma water T_2_ shows potential for addressing that need.

## Data Availability

All data produced in the present study are available upon reasonable request to the authors.

## Acknowledgements

We would like to thank Johnathan Abou-Fadel, Victoria Reid, Ofek Belkin, Mellisa Renteria, Nickolas Sanchez and Charlie Harvey for their efforts and help with this project, as well as Ina Mishra, Clinton Jones, Vipulkumar Patel and Sneha Deodhar for generating data for 44 of the 64 control subjects.

## Author Contributions

Croft, Jacob: Methodology, Writing – Original draft preparation, Writing-reviewing and Editing, Experiments: Data collection, data curation and management; Sandoval, Diana: Writing-Original draft preparations, Experiments-T_2_ data collection and analysis; Cistola, David: Conceptualization, Methodology Writing-Original draft preparation, Reviewing and Editing, Zhang, Jun: Conceptualization, Methodology, Writing-Original draft preparation, Reviewing, Editing, and Finalized manuscript.

## Funding resources

This study was supported by institutional funds from Texas Tech University Health Sciences Center and NIH/NHLBI grant 1 R21 HL143030 to d.p.c.

## Data Availability Statement

R Code script, JMP scripts and data tables are available by request to the authors.

